# The acceptability of testing contacts of confirmed COVID-19 cases using serial, self-administered lateral flow devices as an alternative to self-isolation

**DOI:** 10.1101/2021.03.23.21254168

**Authors:** Nicola Love, Derren Ready, Charlie Turner, Lucy Yardley, G. James Rubin, Susan Hopkins, Isabel Oliver

## Abstract

**Background:** Testing asymptomatic contacts of confirmed COVID-19 cases for the presence of SARS-CoV-2 could reduce onward transmission by improving case ascertainment and lessen the impact of self-isolation on un-infected individuals. This study investigated the feasibility and acceptability of implementing a ‘test to enable approach’ as part of England’s tracing strategy.

**Methods:** Contacts of confirmed COVID-19 cases were offered serial testing as an alternative to self-isolation using daily self-performed lateral flow device (LFD) tests for the first 7 days post exposure. Asymptomatic participants with a negative LFD result were given 24 hours of freedom from self-isolation between each test. A self-collected confirmatory PCR test was performed on testing positive or at the end of the LFD testing period.

**Results:** Of 1,760 contacts, 882 consented to daily testing, with 812 within 48 hours of exposure sent testing packs. Of those who declined to participate, 39.1% stated they had already accessed PCR testing. Of the 812 who were sent packs, 570 (70.2%) reported one or more LFD results; 102 (17.9%) tested positive. Concordance between reported LFD result and a supplied LFD image was 97.1%. 82.8% of PCR positive samples and 99.6% of PCR negative samples were correctly detected by LFD. The proportion of secondary cases from contacts of those who participated in the study and tested positive (6.3%; 95% CI: 3.4-11.1%) were comparable to a comparator group who self-isolated (7.6%; 95% CI: 7.3-7.8%).

**Conclusion:** This study shows a high acceptability, compliance and positivity rates when using self-administered LFDs among contacts of confirmed COVID-19 cases. Offering routine testing as a structured part of the contact tracing process is likely to be an effective method of case ascertainment.

## Introduction

Contacts of confirmed cases of COVID-19 in England are required to isolate for 10 days from the date of their last exposure and are not routinely tested for SARS-CoV-2 unless they develop symptoms (1). Evidence suggests that although people modify their behaviours, full adherence to self-isolation guidance in England may be suboptimal, which may have a detrimental impact on transmission rates (2,3).

Strategies for improving self-isolation compliance have been proposed, including provision of financial or other incentives and penalties for breaking isolation. However, such strategies may not consider the wider economic, social and well-being impacts of isolation (4,5). ‘Test to enable’ strategies are designed to isolate those with infection while reducing or avoiding self-isolation for those without infection. Implementing a ‘test to enable’ approach could therefore lessen the impact of self-isolation on exposed individuals without infection and reduce onward transmission by improving case ascertainment for asymptomatic, paucisymptomatic and pre-symptomatic individuals and, potentially, adherence to self-isolation (6-10). Indeed, a recent study reported that 16.3% of isolating contacts of confirmed cases were PCR positive for SARS-CoV-2 RNA, suggesting that a structureed approach to testing contacts of confirmed cases can support improved case ascertainment (11).

Rapid SARS-CoV-2 antigen testing using lateral flow devices (LFDs) is being increasingly deployed in the UK to screen asymptomatic individuals for SARS-CoV-2 in workplaces, schools and health and social care settings (12). It has been reported that the sensitivity of LFD testing is lower than PCR (57.5-78.8 %); the sensitivity has been reported to improve to >90 % when tested using samples with higher viral loads (13, 14). LFDs provide some added benefits compared to PCR, with a rapid turn-around time (usually less than 30 minutes), low cost and delivery outside of a routine laboratory environment. These features may help increase accessibility to testing for harder to reach groups and allow for a programme of repeated testing, improving the overall case ascertainment (8,9)

Here we describe an investigation into the acceptability of a self-administered daily ‘test to enable’ programme for contacts of confirmed COVID-19 cases using the Innova LFD antigen test during the first 7 days of isolation. On receipt of a negative LFD result, asymptomatic adult participants were exempt from self-isolation for a 24-hour period. The study was designed to identify barriers and strengths to a test to enable approach’ for contacts of confirmed cases of COVID-19, to inform the possible implementation of serial testing of contacts using LFDs as part of England’s NHS Test & Trace strategy.

## Methods

### Recruitment

Asymptomatic adult contacts (> 18 years) exposed to a confirmed COVID-19 case within the preceding 48 hours were identified from NHS Test and Trace (NHS T&T) records and invited to participate by NHS T&T staff using a specific recruitment script (**Figure 1**). Contacts who self-completed contact tracing via a digital route were contacted following the completion of contact tracing, contacts who completed contact tracing via the call centre route were recruited during the routine contact tracing interview. Recruitment was performed Monday to Friday, 8.00 to 16.00, between 11 to 23 December 2020 and 4 to 12 January 2021. A non-probability sample with target size of 800 was sought based on the volume of daily calls performed by recruiters, with no limit on daily recruitment. Individuals contacted by the NHS T&T recruitment team were asked for or their reasons for consenting or declining the testing offer based on predefined categories.

**Figure 1.**
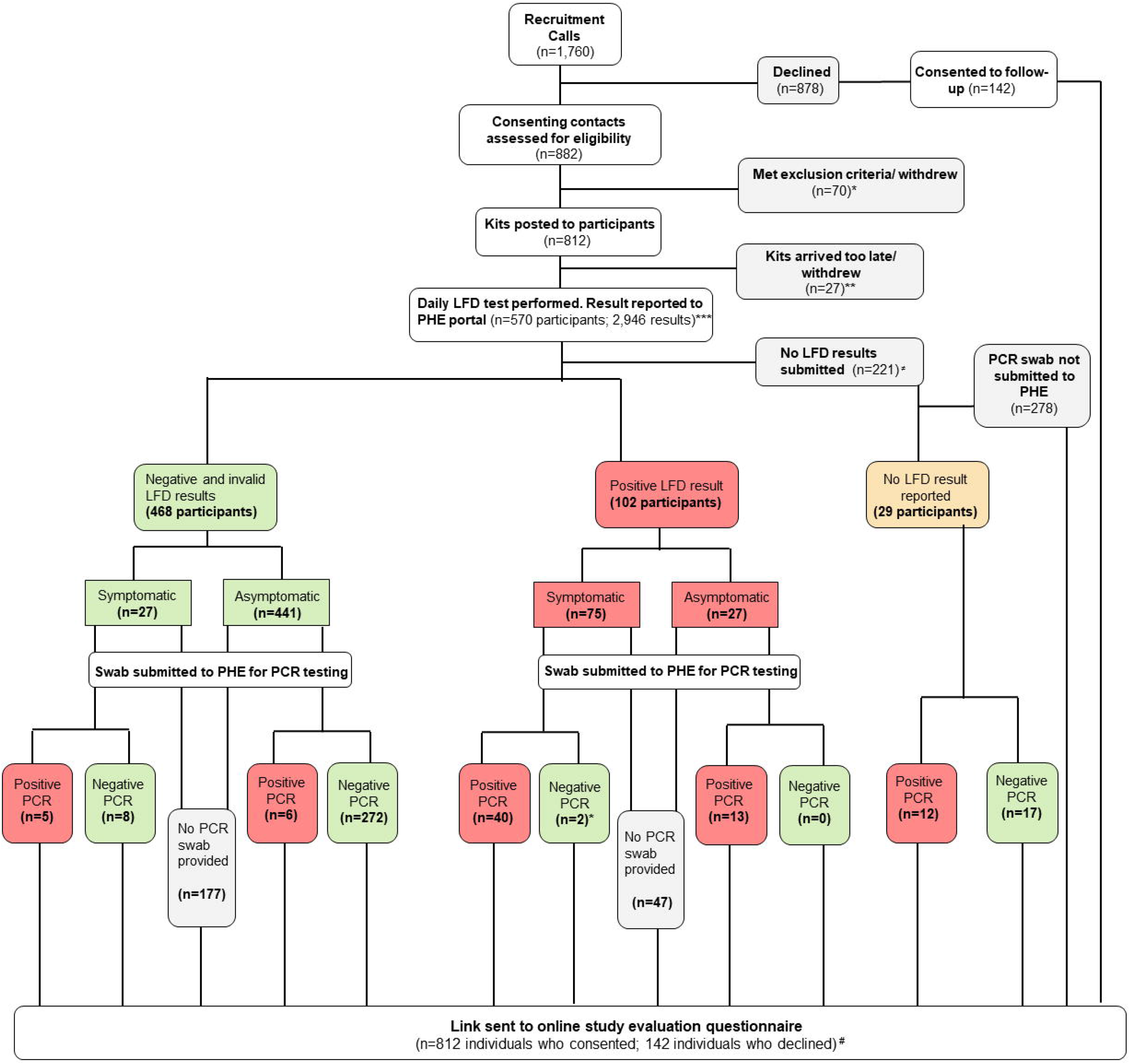
Flow chart of study participation. *68 participants excluded due to exposure >48 hours prior, 1 excluded as non-England resident and 1 withdrew after giving consent. **27 participants notified the study team that their kit arrived too late to participate or did not arrive and were excluded. ***Reported LFD results were excluded if: duplicate entries (n=221; 7.5%), blank entries (n=189; 6.4%), no identifiers (n=27) and reporting by an ineligible non-participant (n=13). ≠5 participants did not report a result but informed the study team that an eligible household member would be reporting using their testing kit. 216 participants did not report a result but did not contact the study team to provide a reason. #Evaluation questionnaire data analysis is presented separately, by Martin et al (15).

### Specimen collection and LFD result reporting

Consenting participants were posted 6 Innova lateral flow devices (LFD; 2 x 3 packs) and a PCR self-sample postal swab kit. Participants were required to self-complete 6 daily LFD tests during the first 7-days post-exposure and self-report daily results together with a digital image of their LFD result to Public Health England (PHE) using a secure online portal developed in Snap Survey, which mirrored the government point of care test portal (www.gov.uk/report-covid-19-result). On receipt of a negative result LFD result, asymptomatic participants were exempt from self-isolation for a 24-hour period.

Self-collected nasal PCR swabs were returned to a PHE accredited laboratory (UKAS ISO 15789) using an approved, trackable postal route on receipt of a positive LFD result or at the end of the 7-day testing period if all results were negative. PCR using an RT-PCR assay evaluated for the detection of SARS-CoV-2 in the ORF1ab assay target and Sarbecoviruses, including SARS-CoV-2 in the E gene target, in clinical respiratory samples was performed on all samples using the ABI QuantStudio 7 Flex real-time PCR system. Reporting of PCR and LFD results was compliant with Health Protection notification regulations and NHS T&T. There were no costs to participants.

### Data linkage

Identifiers from the NHS Test and Trace Contact Tracing and Advice Service (CTAS) webtool were linked to data collected by recruiting agents and to LFD and PCR results. Linkage was deterministic, based on a combination of CTAS ID, name, date of birth, telephone number, postcode and NHS number. Enrichment of NHS number and ethnicity was performed using the NHS demographic batch tracing service (DBS) and Hospital Episode Statistics (HES), respectively. Index of multiple deprivation (IMD) was derived from postcode of residence. PHE’s main laboratory surveillance system, SGSS, and CTAS were interrogated using participant identifiers to obtain non-study PCR test results.

### Data analysis

All data were analysed in Stata version 15. Associations were determined by chi squared and rank sum tests, with a p value less than 0.05 used to show observed differences between groups.

### Secondary attack rates

Secondary attack rates were calculated (in R 4.0.3) for participants who developed COVID-19 as determined by confirmatory PCR, during the study period, using a denominator of all contact case episodes (without deduplication) and a numerator of all case positive contact episodes matched to those contacts with onset within 2-14 days of exposure (without deduplication). A comparator group of all adult cases (with a valid date of birth) reported to NHS T&T who had previously been asymptomatic contacts but were not included in the study, who were exposed to a confirmed case of COVID-19 between 8 – 22 December 2020 or 3–8 January 2021 inclusive was used.

### Ethical considerations

Research governance approval for this study was granted by PHE Research Ethics and Governance Group (REGG) -reference NR0235 on 10 December 2020, with informed consent obtained from participants during recruitment and implied ongoing consent by submission of results or PCR swabs. All data were handled and stored in accordance PHE information governance and securities policies.

## Results

### Study population

1,760 contacts of confirmed cases of COVID-19 were telephoned and asked if they would undertake daily self-testing using LFDs as an alternative to self-isolation; 882 consented to daily contact testing (DCT; 50.1%) and 878 declined (49.9%; **Figure 1**).

The most common self-reported motivations for consenting were a duty to take part (33.8%), the assurance of daily testing (30.4%) and not wanting to self-isolate (26.0%; **Supplementary Table 1**). Reasons for declining were varied, but 39.1% (n=343) declined due to self-reporting a positive PCR result, awaiting a PCR result, or already being part of a routine testing programme (**Supplementary Table 2**). Verifiable PCR results with specimen dates prior to date of interview were obtained for 462 individuals who declined, of which 281 (32.0%) had tested positive between interview and the 14 days prior to exposure (167 who gave this as their reason for declining). Around a third of contacts declined the testing offer citing reasons about the practicalities and commitment of daily testing, with only 1.0% declining due to concerns in LFD performance (n=9).

**Table 1.**
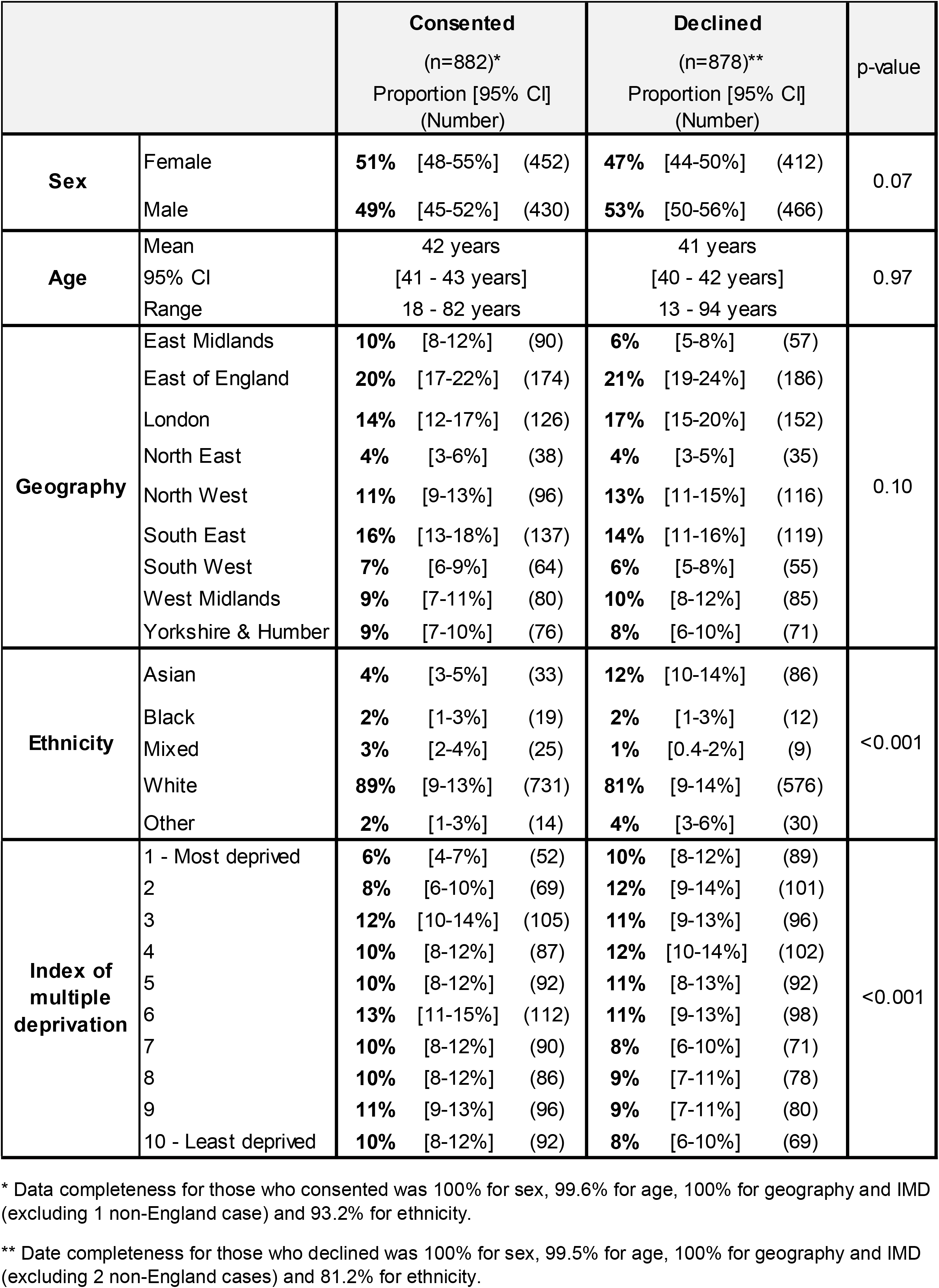
Socio-demographic characteristics of contacts of confirmed cases of COVID-19 (n=1,760) who consented and declined to take part in serial self-testing.

|  |  | **Consented**<br>(n=882)*<br>Proportion [95% CI]<br>(Number) | **Declined**<br>(n=878)**<br>Proportion [95% CI]<br>(Number) | p-value |
| --- | --- | --- | --- | --- |
| **Sex** | Female | **51%** [48-55%] (452) | **47%** [44-50%] (412) | 0.07 |
|  | Male | **49%** [45-52%] (430) | **53%** [50-56%] (466) |  |
| **Age** | Mean | 42 years | 41 years | 0.97 |
|  | 95% CI | [41 - 43 years] | [40 - 42 years] |  |
|  | Range | 18 - 82 years | 13 - 94 years |  |
| **Geography** | East Midlands | **10%** [8-12%] (90) | **6%** [5-8%] (57) | 0.10 |
|  | East of England | **20%** [17-22%] (174) | **21%** [19-24%] (186) |  |
|  | London | **14%** [12-17%] (126) | **17%** [15-20%] (152) |  |
|  | North East | **4%** [3-6%] (38) | **4%** [3-5%] (35) |  |
|  | North West | **11%** [9-13%] (96) | **13%** [11-15%] (116) |  |
|  | South East | **16%** [13-18%] (137) | **14%** [11-16%] (119) |  |
|  | South West | **7%** [6-9%] (64) | **6%** [5-8%] (55) |  |
|  | West Midlands | **9%** [7-11%] (80) | **10%** [8-12%] (85) |  |
|  | Yorkshire & Humber | **9%** [7-10%] (76) | **8%** [6-10%] (71) |  |
| **Ethnicity** | Asian | **4%** [3-5%] (33) | **12%** [10-14%] (86) | <0.001 |
|  | Black | **2%** [1-3%] (19) | **2%** [1-3%] (12) |  |
|  | Mixed | **3%** [2-4%] (25) | **1%** [0.4-2%] (9) |  |
|  | White | **89%** [9-13%] (731) | **81%** [9-14%] (576) |  |
|  | Other | **2%** [1-3%] (14) | **4%** [3-6%] (30) |  |
| **Index of multiple deprivation** | 1 - Most deprived | **6%** [4-7%] (52) | **10%** [8-12%] (89) | <0.001 |
|  | 2 | **8%** [6-10%] (69) | **12%** [9-14%] (101) |  |
|  | 3 | **12%** [10-14%] (105) | **11%** [9-13%] (96) |  |
|  | 4 | **10%** [8-12%] (87) | **12%** [10-14%] (102) |  |
|  | 5 | **10%** [8-12%] (92) | **11%** [8-13%] (92) |  |
|  | 6 | **13%** [11-15%] (112) | **11%** [9-13%] (98) |  |
|  | 7 | **10%** [8-12%] (90) | **8%** [6-10%] (71) |  |
|  | 8 | **10%** [8-12%] (86) | **9%** [7-11%] (78) |  |
|  | 9 | **11%** [9-13%] (96) | **9%** [7-11%] (80) |  |
|  | 10 - Least deprived | **10%** [8-12%] (92) | **8%** [6-10%] (69) |  |

\* Data completeness for those who consented was 100% for sex, 99.6% for age, 100% for geography and IMD (excluding 1 non-England case) and 93.2% for ethnicity.
\*\* Date completeness for those who declined was 100% for sex, 99.5% for age, 100% for geography and IMD (excluding 2 non-England cases) and 81.2% for ethnicity.

**Table 2.**
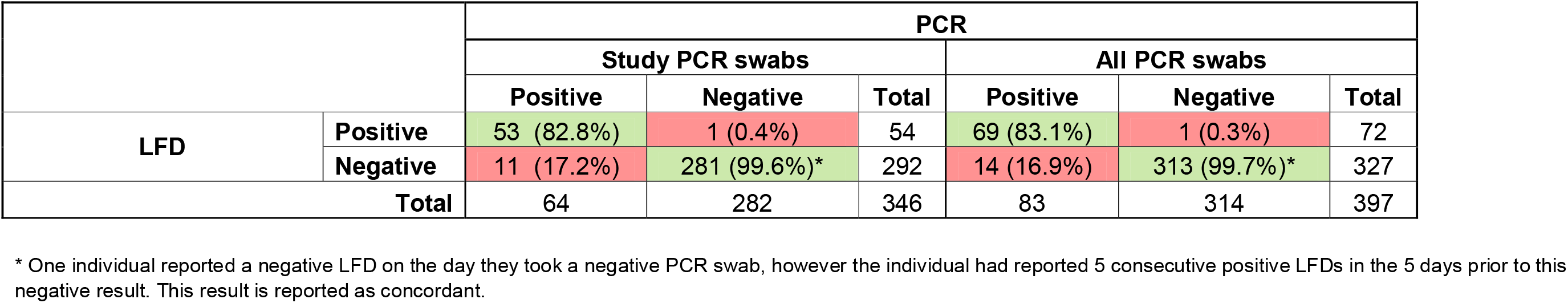
Concordance between LFD and PCR results for individuals who completed a study PCR swab (n=346) and individuals who complated a PCR test via any route (n=51)

There were no significant differences in age, sex or geographical distribution between individuals who consented or declined (**Table 1**). Participants of Asian ethnicity and individuals residing in the two most deprived IMD deciles were more likely to decline, giving observed differences between individuals who consented or declined (p=<0.01). Self-reported motivations for declining participation did not vary significantly between IMD deciles (p=0.267) or ethnic groups (p=0.476; data not shown).

### Timeliness of recruitment and reporting

Contacts recruited after self-completing contact tracing digitally (n=1,553; 88.2%) were selected for inclusion if exposed in the previous 2 days. Those recruited during the contact tracing interview (n=207; 11.8%) were representative of contacts completing the call centre journey, with the median time between exposure and recruitment of 4 days (IQR 2-6 days). Contacts recruited through the call centre journey were more likely to consent to serial testing (68% vs. 48% of those who completed digitally).

812 recruited participants (92%) eligible for inclusion with an exposure within the previous 48 hours were posted a serial testing kit (**Figure 1**; reasons for exclusion given in Figure 1). On average postage took 1.9 days (95% confidence interval: 1.8-2.0 days; n=521), giving a median time between exposure and reporting of first LFD result of 3 days (IQR 2-4 days); for participants recruited after completing the call centre journey this increased to 4 days (IQR 3-6 days).

### LFD results

Between 11 December 2020 and 20 January 2021, 2,946 LFD results were reported, of which 2,505 (85.0%) were eligible for inclusion (**Figure 1**; reasons for exclusion given in Figure 1). 565 of 812 participants who were posted a kit reported at least one LFD result (69.6%), with 5 individuals providing their kit to an eligible family member who reported at least one result and were retrospectively included in the study. Individuals from BAME groups were less likely to report a result after consenting to take part (p=0.01; **Supplementary Table 3**).

**Table 3.**
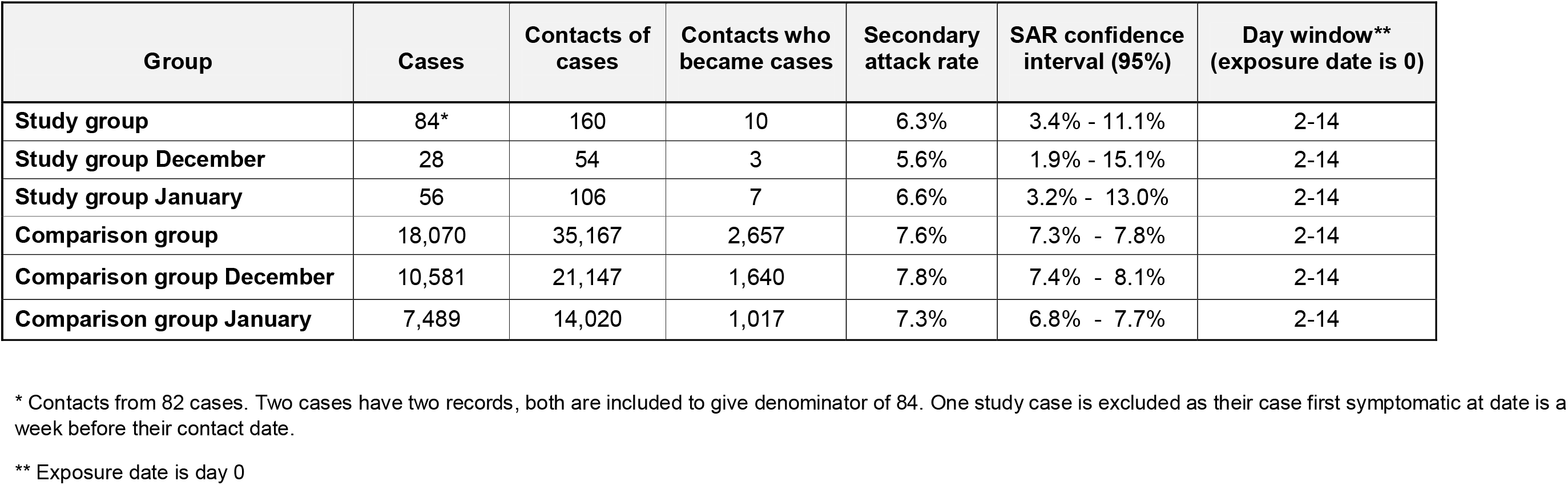
Secondary attack rates for contacts of confirmed cases of COVID-19 who tested positive for SARS-CoV-2 on study PCR swabs (n=84*)

| **Group** | **Cases** | **Contacts of cases** | **Contacts who became cases** | **Secondary attack rate** | **SAR confidence interval (95%)** | **Day window\*\* (exposure date is 0)** |
| --- | --- | --- | --- | --- | --- | --- |
| **Study group** | 84* | 160 | 10 | 6.3% | 3.4% - 11.1% | 2-14 |
| **Study group December** | 28 | 54 | 3 | 5.6% | 1.9% - 15.1% | 2-14 |
| **Study group January** | 56 | 106 | 7 | 6.6% | 3.2% - 13.0% | 2-14 |
| **Comparison group** | 18,070 | 35,167 | 2,657 | 7.6% | 7.3% - 7.8% | 2-14 |
| **Comparison group December** | 10,581 | 21,147 | 1,640 | 7.8% | 7.4% - 8.1% | 2-14 |
| **Comparison group January** | 7,489 | 14,020 | 1,017 | 7.3% | 6.8% - 7.7% | 2-14 |

\* Contacts from 82 cases. Two cases have two records, both are included to give denominator of 84. One study case is excluded as their case first symptomatic at date is a week before their contact date.
\*\* Exposure date is day 0

102 individuals reported at least one positive result (17.9%), 464 (81.4%) reported only negative results and 4 (0.7%) reported only negative and invalid results (**Figure 1**). 75 individuals who tested positive (74%) and 27 who tested negative (6%) self-reported symptoms (13 negative and 6 positive by PCR, 8 no PCR result). A convenience sample of 1,221 LFD images (54.6 % of 2,236 records with an image) were checked by two independent reviewers for concordance with self-reported results (**Supplementary Figure 1)**. 97.1% of images were concordant (n=1,187; 1,132 negative and 55 positive results), 26 images were unreadable (2.1%) and 8 results were discordant (0.8%; 7 positive reported as negative, 1 negative reported as positive). The 7 positive results were from 5 individuals; all were faintly positive, and 3/5 individuals subsequently reported a positive LFD and PCR on receiving a stronger result 1-3 days later.

### Reporting period

The median period from date of exposure to last LFD result reported for individuals testing negative was 8 days (IQR 7-8 days; range 2-16 days). For individuals with a positive LFD result the median period was 5 days (IQR: 3-6; range 2-12 days).Individuals were directed to stop LFD testing on receiving a positive result. The proportion of individuals reporting a positive LFD result decreased with the number of tests a participant performed. 50.6% of individuals who submitted only one LFD result were positive (n=39/77), while only 7 individuals tested positive on their 6^th^ LFD test. 80.1% of the 468 individuals reporting only negative or invalid results (n=375) submitted their final result 7 days after exposure or later.

### Concordance between LFD and self-swab PCR

55/102 participants self-reporting a positive final LFD test (53.9%) returned a study PCR swab; 53 individuals were PCR positive for SARS-CoV-2, of which 37 self-reported becoming symptomatic in the previous 14 days and 15 self-reported remaining asymptomatic on the laboratory request form. Two results were negative; however, one individual reported concordant negative LFD and PCR results after previously testing positive. The median time between reporting a positive LFD result and receipt of PCR swab in the laboratory was 2 days (IQR 1-3 days).

291/468 participants who reported only negative/negative and invalid LFD results returned a PCR swab at the end of the 7-day LFD testing period; 280 individuals were PCR negative (96.2%; n=280/291) and 11 were positive for SARS-CoV-2 (3.8%). 55% of individuals with a positive PCR result and negative LFD result self-reported being symptomatic (n=6/11), compared with 3% of individuals with negative PCR and LFD results (n=9/280). The median time between last negative LFD result and the swab being received at the lab was 3 days (IQR 2-5 days). This gave a concordance between LFD and PCR of 82.8% of PCR positive samples and 99.6% of PCR negative samples correctly detected by LFD for the 61% of participants submitting LFD results and a study PCR swab (**Table 2**).

The median ct (cycle threshold) value for study PCR positive samples from individuals who had tested negative on LFD (n=11) was 24.0 (range 16.9-32.1) for the ORF1ab gene and 25.5 (range 16.9-33.5; one swab negative for ORF1ab) for the E gene target, which was higher than for those individuals who had tested positive on both LFD and PCR (ORF1ab: 20.8 (range: 14.0-35.7); E gene target: 20.0 (range: 13.8-34.2); **Supplementary Table 4**).

SGSS and CTAS systems were also interrogated to determine if individuals requested confirmatory testing through an approved alternative route. 60 individuals who had submitted at least one LFD result obtained a PCR test via an alternative route during their testing period (7 days from exposure) of which 48 were temporally linked to LFD results and were included. Inclusion of these data in the concordance calculation demonstrated 83.1 % of PCR positive samples were detected by LFD and 99.7% of PCR negative samples were correctly reported as negative by LFD for 397 participants with matching data (**Table 2**).

Overall, the PCR positivity for participants who consented to the study and undertook a PCR was 20.9% (83/397), which was comparable to the positivity rate for participants who declined to take part in serial testing but had an identifiable positive PCR result in SGSS or CTAS (20.0%; n=124/620).

### Activities and secondary attack rates

Overall, 160 contacts were reported to NHS T&T from 64 of the 83 participants who tested positive on PCR after submitting at least one LFD result. 19 participants did not report any contacts. Information on activities carried out was also collected for 83 participants. 15.7% reported attending an educational setting, 9.6% a retail setting, 7.2% a healthcare setting, and 41% reported any other workplace setting. Only 2.4% reported a hospitality event and 1.2% a personal care event.

Secondary attack rates (SARs) were calculated for participants who submitted at least one LFD result and received a positive PCR result and compared these to secondary attack rates for a comparator group of all contacts reported to NHS T&T over the same study period who met the same inclusion criteria as study participants, who subsequently became cases. There were no differences in the overall secondary attack rates for the study group (6.3%; 95% CI: 3.4-11.1%) compared with a comparator group (7.6%; 95% CI: 7.3-7.8%). For contacts taking part in the study who tested positive, 96.8% of their contacts were household contacts compared with 94.0% of contacts for cases in the comparator group.

## Discussion

We show daily testing using LFDs was acceptable to contacts of cases of COVID-19 reported to England’s NHS Test & Trace system and that there is likely to be public health benefit in routinely offering tests to contacts to increase case ascertainment in individuals who may not access or who do not qualify for testing under England’s current testing strategy (1).

Uptake of daily testing was encouraging, with 882 (51.1%) of those contacted for recruitment accepting the offer of serial LFD testing. This was higher than previously reported when contacts of cases were offered a PCR swab (39.5%; (11)), which may reflect the additional freedoms awarded by serial testing as part of a test to enable strategy. It is important to note that, in both studies a substantial proportion of those who declined self-reported that they had already been tested for SARS-CoV-2, had booked a test or were part of a testing programme which indicates that testing acceptability is high. Interestingly, 4% of participants declined as they had already tested negative, which is similar to that observed when contacts were offered a PCR swab, being 8.2% (11) and highlights the risk that an early negative result may be providing false re-assurance to individuals who may assume that a negative result will be valid for their full incubation period. Better information on interpretation of negative results may be needed if routine testing of contacts is implemented. Despite controversies in the media about the performance of LFD tests, concerns around test performance or the practicalities of daily testing were not a major issue in this study.

Self-testing kits were sent to 812 individuals by PHE for home LFD testing and PCR validation. Overall, 570 (70.2%) participants returned at least 1 LFD result, however, it is likely that compliance was underestimates as delays to postage, especially around the Christmas period negatively impacted the study. Twenty-seven recruited participants directly contacted the study team to withdraw due to delays in receiving kits, while 65 anonymous respondents reported in the study evaluation questionnaire that they were unable to participate as the kit arrived too late (15). The timeliness of recruitment and kit delivery is a considerable barrier to offering a daily contact testing alternative to self-isolation. It is important to note that individuals submitting to the end of their 7-day period would have been expected to submit fewer than 6 LFD results based on the time lag between exposure and first result. However, many participants completed the 6 LFD tests irrespective of the 7-day testing period.

These data suggest that daily LFD testing was acceptable with high compliance with self-reporting LFD results. However, it is advisable to explore any barriers for individuals who did not to return LFD results to the PHE portal to help improve compliance. Particularly, barriers to uptake need to be further explored in those from Black and minority ethnic (BAME) groups and from lower IMD deciles who were less likely to consent to serial testing and less likely to report a result if posted a test kit, but who are at a higher risk of infection from COVID-19 (16).

102/570 participants reported a positive result via LFD, giving a positivity rate of 17.9%; comparable to that reported when contacts of cases were offered a single PCR swab (16.3%; (11)). Concordance between the Innova LFD and PCR was similar or higher than previously reported for positive results (83.1% vs. 58.9-76.8%) and negative results (99.7% vs. 99.7-99.9%, and higher than previously reported for untrained members of the public self-administering testing (58%; (13,14,17)). Although it’s important to note that a previous study found the differences in sensitivity between expert and non-expert reviewers diminished over time with repeated exposure to performing LFDs, as in serial testing (14,17). LFD failed to detect 11 PCR positive cases of which 5 were symptomatic, it is important to encourage contacts who become symptomatic to access PCR testing. All study PCR swabs were analysed using the same PCR assay, procedures and platforms for those testing negative on LFD but positive on PCR, median Ct values were higher by 3.2-5.5 cycles. Other studies have noted that Ct values are an important determinant for detection of SARS-CoV-2 (18).

Self-testing is trust based and puts an onus on the contact to perform the test correctly and report the result. Although only a sub-set of participants did a confirmatory PCR, we observed good concordance suggesting tests were being performed correctly. Furthermore, blinded analysis of images by two reviewers found just 8 discordant results reported from 6 individuals. One result was clearly negative and may have been misreported as positive. The 7 positive results reported as negative were very faintly positive and instructions for contacts may need to be made clearer to ensure accurate interpretation of results.

## Conclusion

This study shows a high acceptability and compliance with self-testing among many contacts of confirmed COVID-19 cases. Therefore, offering routine testing as part of the contact tracing process is likely to be an effective method of case finding and daily testing with release from self-isolation if negative, may improve adherence with self-isolation. Offering a structured programme of testing of contacts of cases still requires some additional questions to be addressed, specifically, which testing strategy is most effective. Further work is needed to understand any impact of daily testing on secondary transmission.

## Limitations of the study

Sample size was not determined by statistical power considerations and this is an important limitation that should be considered when interpreting the findings. The study was designed to investigate the feasibility and acceptability of a test to enable study, with further research planned to use more robust methodologies to look at associations. Secondary attack rates in particular should be interpreted with caution due to small numbers, with estimates of uncertainty for secondary attack statistics subject to random variation when underlying cohorts are small. Only close contacts who are named by the case are captured by contact tracing. These attack rates should be considered minimum estimates both because only those contacts who access testing can subsequently be identified as a case and because the matching process used to detect when a contact goes onto become case within NHS Test and Trace is highly specific. Secondary attack rates are also likely to underestimate the number of contacts, given the predominance towards the reporting of household contacts, which may be a result of the study being performed during a period of lockdown in England. A large proportion of contacts who became cases had reported activities outside of the household during their infectious period to NHS T&T but declared no contacts in these settings to NHS T&T.

## Data Availability

No additional data available

## Acknowledgements

The study team would like to thank the participants of this study for their time and participation. Acknowledgements also go to PHE data entry staff, PHE laboratory staff, PHE CTAS team, PHE Health Intelligence Division, Koren Jones and Áine Kiernan from the SGSS team and DH, NHS T&T and PHE staff consulted in the design of this study. Particular thanks go to Joan Henderson, Marina Vabistsevits and Cong Chen who provided support with CTAS data and secondary attack rates, and to Simon Cockburn, Gillian Atkinson, Hazel Coulson, Katie Fuller, Taran Huxtable and the Agile Lighthouse team.

