## Supplementary Figures for "The acceptability of testing contacts of confirmed COVID-19 cases using serial, self-administered lateral flow devices as an alternative to self-isolation"

**Supplementary Table 1 – Self-reported reasons for consenting to take part in the serial self-testing study**

| Reason for consenting | Number of participants | Percentage of participants |
|---|---|---|
| Duty to take part/societal benefit | 298 | 33.8% |
| Would like the assurance of daily testing | 268 | 30.4% |
| Do not wish to isolate for 10/14 days | 229 | 26.0% |
| Other - unspecified reason/ no reason given | 58 | 6.6% |
| Convenience e.g. shopping, exercise | 16 | 1.8% |
| To return to work | 9 | 1.0% |
| Curiosity | 2 | 0.2% |
| Other - specified reason | 2 | 0.2% |
| **Total** | **882** | **100%** |

**Supplementary Table 2 – Self-reported reasons for declining to take part in the serial self-testing study**

| Reason for Declining | Number of participants | Percentage of participants |
|---|---|---|
| Already tested positive | 197 | 22.4% |
| Aready tested and awaiting result | 127 | 14.5% |
| Does not want to commit to daily testing | 102 | 11.6% |
| Does not want to perform self-testing | 69 | 7.9% |
| Would prefer to self isolate/made arrangements to isolate | 56 | 6.4% |
| Uncooperative | 45 | 5.1% |
| Already booked a test | 33 | 3.8% |
| Already tested negative | 33 | 3.8% |
| Other - specified reason | 21 | 2.4% |
| Does not want to waste resources | 20 | 2.3% |
| Already part of a testing programme | 19 | 2.2% |
| Household member is a case | 19 | 2.2% |
| Met exclusion criteria | 17 | 1.9% |
| Would rather isolate with rest of household | 16 | 1.8% |
| Did not want to isolate for longer if testing positive | 15 | 1.7% |
| Length of time for testing is too long | 15 | 1.7% |
| Not willing to take part in research | 15 | 1.7% |
| Is unable to perform self-testing | 11 | 1.3% |
| Other - unspecified reason/ no reason given | 11 | 1.3% |
| At end of isolation period | 9 | 1.0% |
| Concerns about test performance | 9 | 1.0% |
| Concerns about spreading infection | 5 | 0.6% |
| Unable to work so prefer to isolate | 5 | 0.6% |
| Language barrier | 4 | 0.5% |
| Would find testing uncomfortable | 3 | 0.3% |
| Technological barrier | 2 | 0.2% |
| **Total** | **878** | **100%** |

**Supplementary Table 3 - Socio-demographic characteristics of contacts of confirmed cases of COVID-19 who consented and reported a result (n=570*) and those posted a kit who did not report a result (n=220**)**

| | | Reported (n=570*) Proportion (Number) | | | Did not report (n=220**) Proportion (Number) | | | p-value |
|---|---|---|---|---|---|---|---|---|
| **Sex** | Female | 51% | [47-55%] | (291) | 50% | [43-56%] | (110) | 0.79 |
| | Male | 49% | [45-53%] | (279) | 50% | [44-57%] | (110) | |
| **Age** | Mean | | 42 | | | 42 | | 0.77 |
| | 95% CI | | 41 - 43 | | | 40 - 44 | | |
| | Range | | 18 - 80 | | | 18 - 80 | | |
| **Geography** | East Midlands | 11% | [8-14%] | (63) | 9% | [5-12%] | (19) | 0.45 |
| | East of England | 21% | [18-25%] | (121) | 16% | [11-21%] | (36) | |
| | London | 12% | [9-15%] | (68) | 16% | [11-21%] | (36) | |
| | North East | 5% | [3-6%] | (26) | 5% | [2-7%] | (10) | |
| | North West | 12% | [9-14%] | (66) | 9% | [5-13%] | (20) | |
| | South East | 16% | [13-19%] | (89) | 16% | [11-21%] | (36) | |
| | South West | 7% | [5-9%] | (40) | 9% | [5-12%] | (19) | |
| | West Midlands | 8% | [6-11%] | (48) | 11% | [7-15%] | (24) | |
| | Yorkshire & Humber | 9% | [6-11%] | (49) | 9% | [5-13%] | (20) | |
| **Ethnicity** | Asian | 3% | [2-4%] | (16) | 6% | [3-9%] | (12) | 0.01 |
| | Black | 2% | [0.5-3%] | (8) | 5% | [2-7%] | (9) | |
| | Mixed | 4% | [2-6%] | (22) | 2% | [0-3%] | (3) | |
| | White | 90% | [88-93%] | (481) | 85% | [81-90%] | (170) | |
| | Other | 1% | [0.2-2%] | (6) | 3% | [0.3-5%] | (5) | |
| **Index of multiple deprivation** | 1 - Most deprived | 5% | [4-7%] | (31) | 5% | [2-8%] | (12) | 0.33 |
| | 2 | 7% | [5-9%] | (41) | 10% | [6-13%] | (21) | |
| | 3 | 11% | [9-14%] | (65) | 14% | [9-19%] | (31) | |
| | 4 | 10% | [8-13%] | (58) | 10% | [6-14%] | (23) | |
| | 5 | 9% | [6-11%] | (50) | 11% | [7-16%] | (25) | |
| | 6 | 15% | [12-18%] | (84) | 9% | [5-13%] | (19) | |
| | 7 | 9% | [7-11%] | (51) | 10% | [6-15%] | (23) | |
| | 8 | 12% | [9-14%] | (66) | 7% | [3-10%] | (15) | |
| | 9 | 12% | [9-14%] | (66) | 10% | [6-13%] | (21) | |
| | 10 - Least deprived | 10% | [8-13%] | (58) | 14% | [9-18%] | (30) | |

* Includes 5 participants who were not recruited via the Agile Lighthouse. Data completeness for those who consented was 100% for sex, 99.6% for age, 100% for geography and IMD (excluding 1 non-England case) and 93.5% for ethnicity.

** Excludes 27 of 247 participants who did not report an LFD result but were known to have not received a kit/received a kit too late to participate. Date completeness for those who declined was 100% for sex, 99.5% for age, 100% for geography and IMD and 90.4% for ethnicity.

**Supplementary Figure 1 – Participant images showing range of positive results obtained**

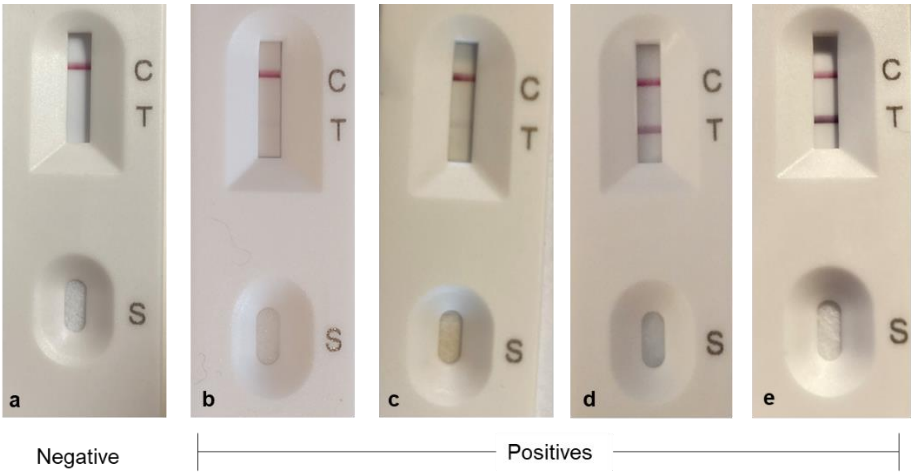

**Supplementary Table 4 – CT Values obtained for positive samples tested at PHE Colindale, individuals reporting at least one LFD result prior to PCR swab**

| ct (cycle threshold value) | All (n=64) | | Positive LFD positive PCR (n=53) | | Negative LFD positive PCR (n=11) | |
|---|---|---|---|---|---|---|
| | ORF1ab* | E gene | ORF1ab | E gene | ORF1ab* | E gene |
| Mean | 22.8 | 22.4 | 22.3 | 22.0 | 24.0 | 24.6 |
| Median | 21.9 | 21.0 | 20.8 | 20.0 | 24.0 | 25.5 |
| Min | 14.0 | 13.8 | 14.0 | 13.8 | 16.9 | 16.9 |
| Max | 35.7 | 34.2 | 35.7 | 34.2 | 32.1 | 33.5 |

*One PCR sample was negative for the ORF1ab gene target
