## Supplementary Methods for "The acceptability of testing contacts of confirmed COVID-19 cases using serial, self-administered lateral flow devices as an alternative to self-isolation"

### Study population and recruitment

Asymptomatic adult contacts of confirmed COVID-19 cases were invited to participate by NHS T&T Tier 3 Agile Lighthouse call handlers. Individuals were identified following self-completion or household contact completion of the NHS T&T contact tracing questionnaire. Contacts were contacted and invited to participate following completion of the NHS T&T digital journey if their date of last exposure to the case had taken place within the previous 48 hours. In addition, contacts completing the 'call centre journey' were recruited during the routine contact tracing interview. A flow diagram of recruitment and reporting processes can be found in Supplementary methods Figure 1.

Recruitment was performed Monday to Friday, 8.00 to 16.00, from 11 to 23 December 2020 and from 4 to 12 January 2021, with no limit on the daily number of participants recruited. A non-probability sample with target size of 800 was sought based on the volume of Agile Lighthouse daily calls over a 14-day period.

Consenting participants were posted a study testing pack consisting of 6 lateral flow devices (LFD; 2 x 3 packs), a PCR self-sample postal swab kit and study documentation. Participants recruited before 14.30 each day were sent a kit by Royal Mail the same day. A text message was sent to all participants with a valid mobile number to inform them that their kit had been sent, and to provide participants with the hyperlink to the results portal, using the Notify messaging service (https://www.notifications.service.gov.uk).

Confirmatory self-collected swabs were returned to the UKAS (ISO 15789) accredited laboratory at PHE Colindale for PCR validation and quality assurance of LFD testing results, using an approved Royal Mail route and bespoke test request form for this study. The PCR assay was carried out by an independent team who were masked from the LFD results. Swabs were tested using an RT-PCR assay designed and evaluated for the detection of SARS-CoV-2 in clinical respiratory samples, using the ABI QuantStudio 7 Flex real-time PCR system. The assay specifically detected SARS-CoV-2 in the Orf1ab assay target, and Sarbecoviruses including SARS CoV-2 in the E gene target. The assay was intended to be performed for diagnosis of COVID-19 infection, and for confirmation of SARS-CoV-2 LFD results identified using the Innova LFD assay. All PCR consumable and processing costs plus postage costs were met by Public Health England (PHE).

### Specimen collection and result reporting

Consenting individuals were asked to self-complete up to 6 daily LFD tests and self-report results to PHE each day using a secure results portal developed in Snap Survey. The portal mirrored the government point of care test portal (www.gov.uk/report-covid-19-result), which was not operational when the study commenced. Participants were required to submit daily LFD results, an image of the test, identifiers (name, date of birth, postcode, NHS number), symptoms and the date the kit arrived.

Results were uploaded to the central PHE point of care testing (POCT) result portal to ensure notification was compliant with the regulations that state all COVID-19 positive, indeterminate, negative and void results are reported to Public Health England (PHE) as set out in amendments to The Health Protection (Notification) Regulations 2010.

Notify text messaging reminders were sent to encourage individuals to submit a confirmatory home collected PCR swab on submission of a positive LFD result or at the end of their 7-day LFD testing period. Confirmatory PCR test results were carried out a PHE Colindale and results were uploaded to The PHE Modular Open Laboratory Information System (MOLIS)

**Supplementary methods Figure 1– Flow diagram of recruitment, study participant flow and reporting processes.**

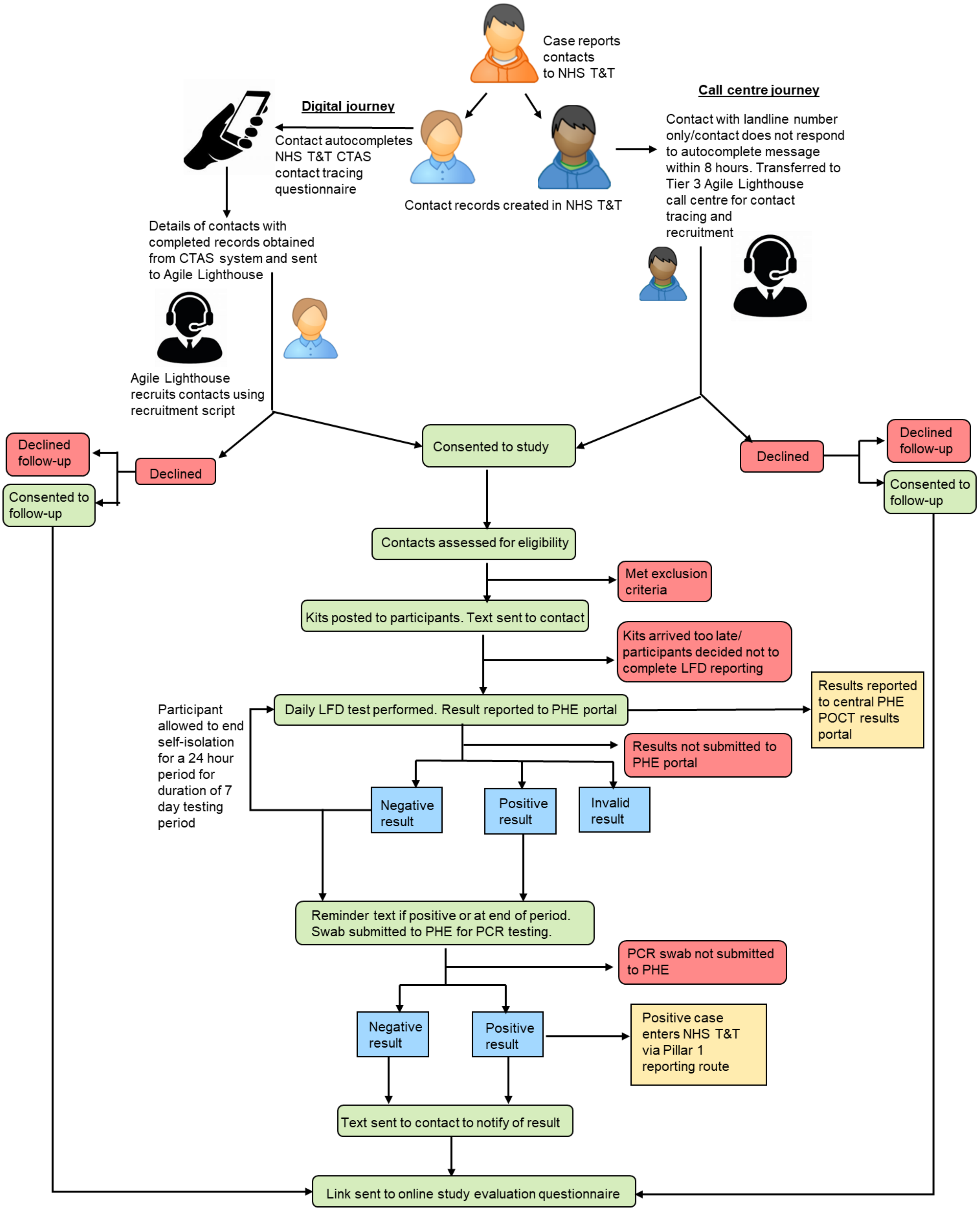

with study samples identifiable using a study specific requestor code (NISFS2). Participants were notified of their PCR result using the Notify text message system. Individuals with a positive PCR result were referred to NHS Test and Trace using the standard laboratory reporting processes via the Second-Generation Surveillance System (SGSS).

#### Study evaluation

A link to an electronic evaluation questionnaire developed in Snap Survey was sent by Notify text messaging to all consenting participants and all declining participants, who consented to contact from academic partners, if they had a mobile number. A third group of individuals, who were contacts completing a routine self-isolation NHS T&T journey over the same time period, who were eligible for inclusion but not offered serial testing and who had agreed to further contact from NHS T&T were also sent a link and reminder text to the evaluation questionnaire. The questionnaire covered reasons for consenting and declining, confidence in LFD results, issues in conducting self-testing, activities undertaken when testing negative by LFD and when self-isolating, contact reported when testing negative and when self-isolating, and demographic characteristics. Questions asked to each of the three groups were dependent on participation status.

#### Data sources and data linkage

Demographic data from the NHS Test and Trace Contact Tracing and Advice Service (CTAS) webtool (name, date of birth, email address, mobile number, sex and NHS number) was linked to recruitment questionnaire data obtained by the Agile Lighthouse during participant recruitment (address including postcode, ethnicity, consent, reason for accepting/declining) using the participants unique CTAS ID. Missing NHS numbers were obtained from the NHS demographic batch tracing service (DBS), with NHS number linked to Hospital Episode Statistics (HES) data to improve ethnicity completeness.

Participants were asked to submit daily LFD test results to the results portal, together with an image of the LFD test result and identifiers to enable linkage to other data sources (name, date of birth, postcode, NHS number). Participants were also asked to provide the date their test kit arrived and to state if they had symptoms prior to taking the test.

Demographic data from the NHS Test and Trace Contact Tracing and Advice Service (CTAS) webtool collected during completion of the NHS T&T contact questionnaire (name, date of birth, email address, mobile number, sex and NHS number) was deterministically linked to recruitment questionnaire data obtained by the Agile Lighthouse (address including postcode, ethnicity, consent, reason for accepting/declining) using the participants CTAS ID. Results submitted to the LFD portal were deterministically linked to this demographic and recruitment data and to PCR results obtained from MOLIS, which included demographic information and symptomatology recorded on the laboratory request form sent with the confirmatory PCR swab. Linkage of datasets was deterministic, based on the inclusion of a combination of CTAS ID, name, date of birth, telephone number, postcode and NHS number in all data sets.

Deterministic linkage using NHS Number, name, date of birth and/or postcode was also used to identify any contacted individuals (both consenting and declining inclusion) in PHEs Second Generation Surveillance System (SGSS) and name and telephone number was used to check for any case records in the CTAS system. This was to determine if individuals contacted in this study had obtained SARS-CoV-2 testing via another route during the study period.

### Data analysis

All data was analysed as of 8 February 2021. Following linkage, the dataset was analysed in Stata version 15. Associations were determined by Chi2 and rank sum test, with a p value of <0.05 used to show significant observed differences between groups. Sample size was not determined by statistical power considerations and this should be considered when interpreting study results.

This study was not designed to estimate secondary attack rates due to small numbers. However, secondary attack rates were calculated using the denominator of all contact case episodes identified (no deduplication if a person was named twice by the same case or a different one) and the numerator of all case positive contact episodes matched to those contacts (no deduplication if a person became a case twice in our dataset within a relevant period. The comparison group was made up of cases who had previously been contacts, who did not report any symptoms as contacts (note that symptom data for contacts is often poorly completed and in some situations it is not possible for contacts to provide that information), who were over 18 in any of their contact/case age data provided, who were exposed as contacts in the periods 2020-12-08 – 2020-12-22 or 2021-01-03 – 2021-01-08 and were not in the study group.

### Ethical considerations

Research governance approval for this study was granted by PHE Research Ethics and Governance Group (REGG) - reference NR0235. All data were handled and stored in accordance with the Data Protection Act (2018) and GDPR in line with PHE information governance and securities policies.

Informed consent for both serial testing and evaluation interviews with academic partners was obtained by Tier 3 Agile lighthouse staff during the recruitment call. Implied consent of participants was assumed inferred by their return of the self-test LFD results, collection and return of the self-test PCR swab and completion of the accompanying laboratory request form. Individuals who did not provided consent, either actively or because they lacked capacity were not included in the study.
